# Loneliness and diurnal cortisol levels during COVID-19 lockdown: the roles of living situation, relationship status and relationship quality

**DOI:** 10.1101/2022.02.25.22271461

**Authors:** Dora Hopf, Ekaterina Schneider, Corina Aguilar-Raab, Dirk Scheele, Beate Ditzen, Monika Eckstein

**Author notes:** shared senior author position (Beate Ditzen, Monika Eckstein).

## Abstract

Loneliness and social isolation have become increasing concerns during COVID-19 lockdown through neuroendocrine stress-reactions, physical and mental health problems. We investigated living situation, relationship status and quality as potential moderators for trait and state loneliness and salivary cortisol levels (hormonal stress-responses) in healthy adults during the first lockdown in Germany. *N*=1242 participants (mean age = 36.32, 78% female) filled out an online questionnaire on demographics, trait loneliness and relationship quality. Next, *N*=247 (mean age = 32.6, 70% female) completed ecological momentary assessment (EMA), collecting twelve saliva samples on two days and simultaneously reporting their momentary loneliness levels. Divorced/widowed showed highest trait loneliness, followed by singles and partnerships. The latter displayed lower momentary loneliness and cortisol levels compared to singles. Relationship satisfaction significantly reduced loneliness levels in participants with a partner and those who were living apart from their partner reported loneliness levels similar to singles living alone. Living alone was associated with lower loneliness levels. Hierarchical linear models revealed a significant cross-level interaction between relationship status and momentary loneliness in predicting cortisol. The results imply that widowhood, being single, living alone and low relationship quality represent risk factors for loneliness and having a partner buffers neuroendocrine stress responses during lockdown.

## Introduction

The recent Corona virus pandemic (COVID-19) has been occupying mental and physical health facilities for two years now. Hard lockdown regulations in almost all countries early during the pandemic (April until June 2020) to prevent further spreading of the virus entail increased social isolation. The steady and massive health threat from the virus in combination with the missing social buffering effect of everyday social encounters lead to or amplified psychosocial problems that could have long-term consequences for mental and physical health^1-4^. E.g., loneliness, as the subjective and emotional component of social exclusion, is a highly topical and public health issue in modern societies, where social isolation and anonymity become increasingly prevalent^5,6^. It entails the perceived lack of intimacy or social companionship and the feeling that social relationships are deficient in either quality or quantity^7^. By contrast, social isolation is defined as the objective state of being alone^7,8^. According to the belongingness-hypothesis, loneliness is rooted in the human need to socially belong, or the pervasive drive to form and maintain lasting positive and significant social relationships^9^. It has been shown that the sense of belonging in early adolescents is mainly achieved through the acceptance by peers, whereas in late adolescence and adulthood, it is achieved especially by romantic relationships, marital status and close friends^10^. On the other hand, lacking feelings of belonging are assumed to be associated with loneliness and negative physical and mental health outcomes in a long-term^9^. Both loneliness and social isolation are significantly associated with indices of physical and mental health, such as psychosocial stress^11^, depression^12^, generalized anxiety^6^, cardiovascular diseases^13^, chronic obstructive pulmonary disease^14^, and mortality^8,15-19^. Chronic loneliness may hamper the formation of new social relationships by inducing negative cognitive biases such as interpersonal distrust^20^. Furthermore, loneliness is associated with neuroendocrine parameters, like elevated cortisol levels^21-23^ and altered cortisol awakening responses^23,24^. As one of the main effector hormones of the hypothalamus-pituitary-adrenal (HPA) axis, the steroid cortisol is secreted in response to external and internal stressors in order to re-establish homeostasis^25^. Previous studies suggest that cortisol may serve as a potential short-term correlate of loneliness, predicting poor physical or mental health outcomes in the long-term^21,22^.

According to the social buffering hypothesis^26^, social relationships play a beneficial role in physical and mental health^26-29^. Among the most intense social relationships are romantic relationships, as they serve as the primary source of support, fulfilling needs such as intimacy, attachment, and emotional support^30^. Supportive and affectionate interactions with the partner reduce stress, pain, and psychological distress. They even influence the immune system, wound healing or mortality rates^31-35^. Being in a relationship has been found to be associated with lower loneliness levels, compared to never-married, divorced, and widowed individuals^36-38^. Especially in the middle and higher age, romantic relationships become important buffers for loneliness^39^. Furthermore, romantic relationships directly affect physiological stress responses, such as cortisol secretion. Individuals who are in a close relationship, show lower aggregated cortisol levels than singles^40^ and affectionate couple interaction can reduce cortisol levels^41,42^. On the other hand, the loss of a partner, for example due to breakup or death, is considered one of the most stressful life events in adulthood, being associated with reduced mental and physical health outcomes^43^. Divorced and widowed individuals show significantly higher loneliness scores than married individuals^44-46^. Furthermore, partner loss is accompanied by altered HPA axis functioning, resulting in elevated cortisol levels and flattened diurnal cortisol slopes^47^.

Although being in a relationship protects against feelings of loneliness, couples can also experience higher levels of loneliness. This can be explained by the cognitive perspective^48^, according to which loneliness is further influenced by the quality and not only the quantity of social relationships. As one important factor, relationship quality has been shown to be negatively associated with loneliness,^49-53^. In times of extreme social isolation, relationship quality might become an important moderator, especially if couples do not live together and thus are unable to see their partner and potentially have to rely on non-physical relationship qualities. Living alone has become increasingly prevalent, with one-person households accounting for more than 40% of all households in Scandinavian nations, more than 33% of all households in France, Germany, and England; and more than 25% of all households in the United States, Russia, Canada, Spain, and Japan^54^. In Germany, in the young adult age of 18 to 30 years, more than 30% live without a partner^55^. An important distinction in this context is between partnerships with and without a common household (the latter being called “living apart together”). In general, living alone has been seen as a risk factor for poor physical and mental health^54,56^. For instance, the living situation predicts mortality risk^57,58^ and people who are living alone show higher loneliness levels^59^.Cross-sectional studies suggest that during the pandemic, being married served as a protective factor against loneliness^60^, whereas being divorced or widowed increased the risk of loneliness^61^. Furthermore, living with others has been found to protect against loneliness^62^, even when controlling for relationship status^63^ and loneliness during lockdown predicted psychological distress^64^. It has not been investigated yet, however, whether relationship status and living situation during lockdown affected biological, specifically neuroendocrine, health parameters, such as cortisol levels. In previous studies, living alone had been positively associated with cortisol levels^65^. Furthermore, the buffering effect of living situation and relationship status with regard to psychobiological outcomes during stress-exposure (i.e. the world-wide considerable psychological stress through COVID-19) has not been investigated yet. Previous research suggests that the separation from a partner is associated with elevated feelings of loneliness and cortisol levels in general^66-68^. In adolescents, significant correlations between self-reported loneliness and cortisol awakening responses during COVID-19 lockdown were found^69^. However, moment-to-moment associations of loneliness and cortisol have not been investigated in adults yet. Furthermore, it is still elusive if relationship status and living situation moderate these associations. Lastly, the effect of psychological variables such as relationship satisfaction, on the association between living arrangements and loneliness during lockdown has not yet been addressed.

### Study objectives

The purpose of this study was to investigate relationship status and living situation as potential moderators for trait and state loneliness as well as momentary cortisol levels during the COVID-19 pandemic and during lockdown. We aimed to replicate findings about the association between relationship status and trait loneliness (Hypothesis 1). In order to explore state loneliness and cortisol in every-day life, we used an ecological momentary assessment (EMA) approach. Secondly, we expected that the current living situation and relationship status have an impact on momentary (state) loneliness (Hypothesis 2) and cortisol levels (Hypothesis 3). Based on previous studies, we assumed, that both relationship status and living situation would have independent effects on loneliness and cortisol. Furthermore, we hypothesized a positive association between momentary (state) loneliness and momentary (state) cortisol levels (Hypothesis 4) and expected that relationship status and living situation moderate this association (Hypothesis 5). Lastly, we hypothesized that relationship quality would moderate the association between living situation and momentary (state) loneliness levels in individuals being in a relationship (Hypothesis 6).

## Methods

### Participants

This study was approved by the Heidelberg Medical Faculty’s Ethics Committee (Heidelberg University, approval no. S-214/2020) and registered online (https://www.drks.de/drks_web/navigate.do?navigationId=trial.HTML&TRIAL_ID=DRKS00021671). It was performed in accordance with the Declaration of Helsinki. All participants signed an informed consent and were recruited between April 1^st^ and July 30^th^ 2020 via online media and local newspapers. Inclusion criteria were: Fluency in German, minimum age of 18 years and willingness to participate voluntarily. In total, 1483 individuals agreed to participate, from which 1054 participants filled out the questionnaires of interest (see Figure 1). The mean age of the participants was *M* = 36.32 years (*SD* = 14.75, *Range* = 18;81), with 77.7 % being female (*n* = 819). Demographic characteristics are displayed in Table 1.

**Table 1.** Demographic characteristics of study 1 (online survey).

|  | **Categories** | ***n* (%)** |
| --- | --- | --- |
| Gender | Female | 819 (77.7) |
|  | Male | 227 (21.5) |
|  | Diverse | 4 ( .4) |
|  | Non-responders | 4 ( .4) |
| Occupation | At school/training/college/university | 368 (34.9) |
|  | Employed/civil servant | 502 (47.6) |
|  | Self-employed | 100 (9.5) |
|  | Unemployed | 40 (3.8) |
|  | Pensioner/Housewife/househusband | 98 (9.3) |
| Relationship status | In a relationship | 655 (77.7) |
|  | Single | 329 (31.2) |
|  | Divorced/widowed | 70 (6.6) |

**Figure 1.**
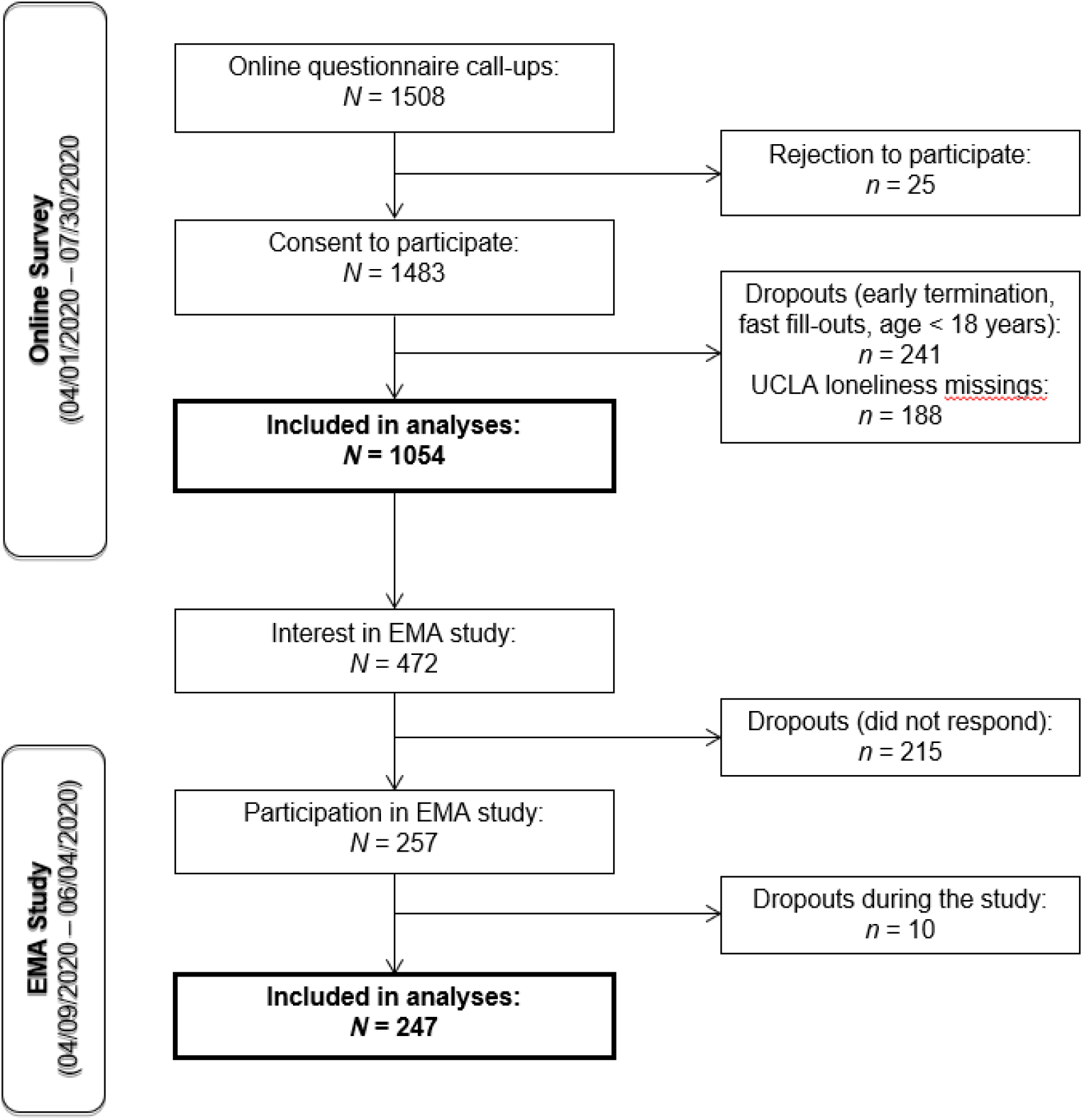
Flowchart of the recruitment process.

Of the participants in the online survey, 472 showed interest in the EMA with the salivary sampling. Of those 472 participants, 54% (*n* = 257) took part in the EMA study. After excluding individuals who did not react to our messages and dropouts during data collection (*n* = 10), the remaining 247 cases were included in the analyses. The participants’ mean age was *M* = 32.6 years (*SD* = 13.12, *Range* = 18;78), with 70 % being female (*n* = 173). Demographic characteristics of the EMA study sample are displayed in Table 2.

**Table 2.**
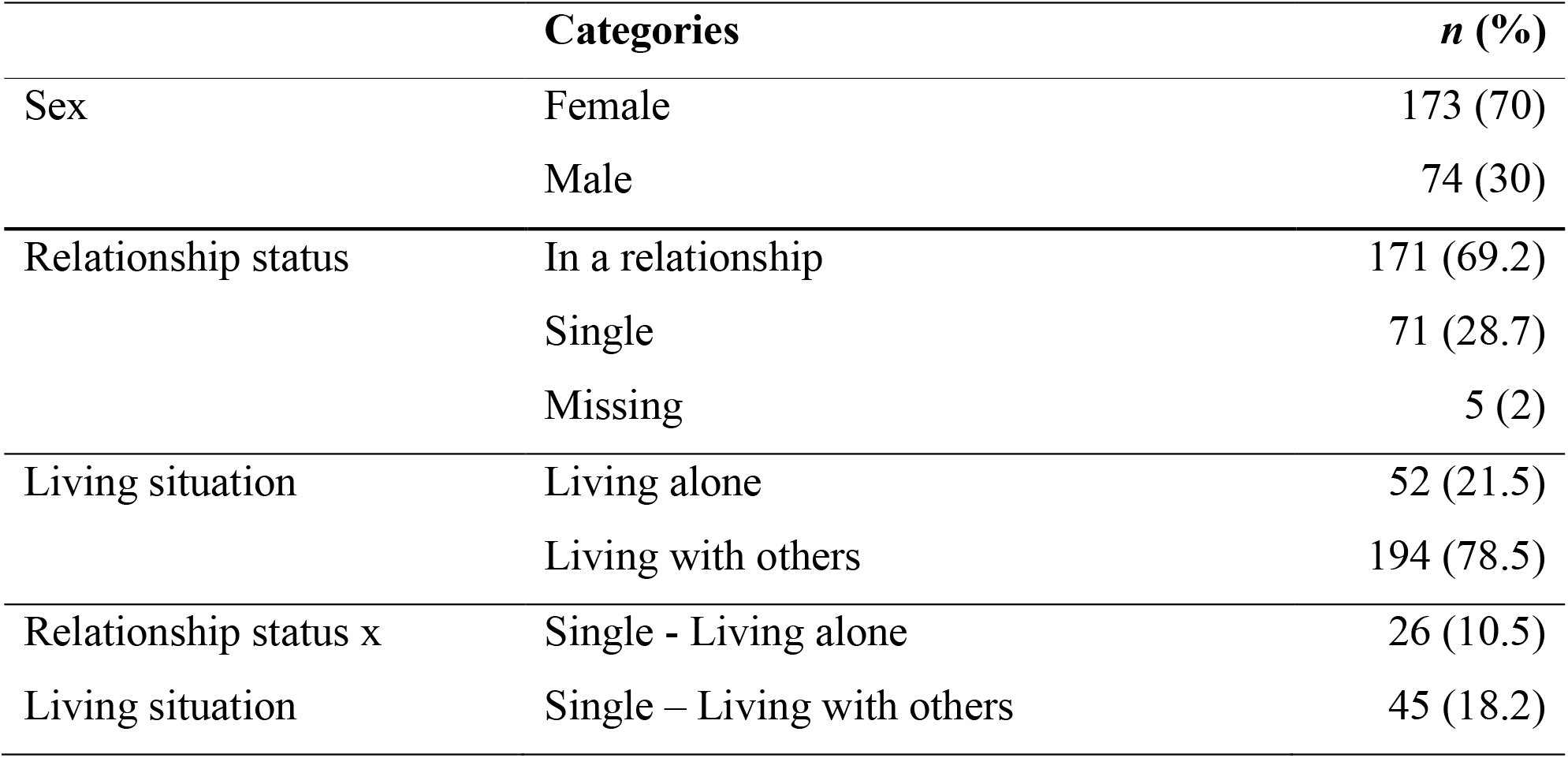
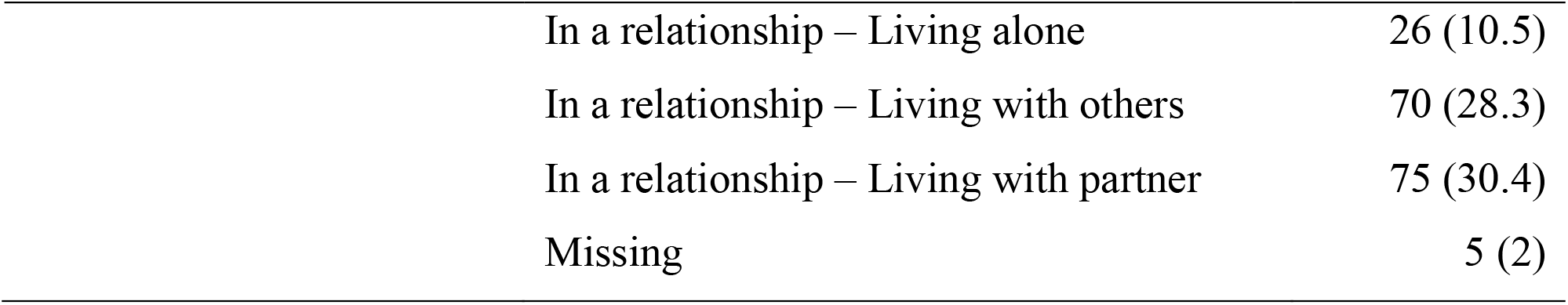
Demographic characteristics of the EMA study.

|  | **Categories** | ***n* (%)** |
| --- | --- | --- |
| Sex | Female | 173 (70) |
|  | Male | 74 (30) |
| Relationship status | In a relationship | 171 (69.2) |
|  | Single | 71 (28.7) |
|  | Missing | 5 (2) |
| Living situation | Living alone | 52 (21.5) |
|  | Living with others | 194 (78.5) |
| Relationship status x | Single - Living alone | 26 (10.5) |
| Living situation | Single – Living with others | 45 (18.2) |

|  |  |
| --- | --- |
| In a relationship – Living alone | 26 (10.5) |
| In a relationship – Living with others | 70 (28.3) |
| In a relationship – Living with partner | 75 (30.4) |
| Missing | 5 (2) |

### Measures

#### Loneliness

To measure trait loneliness in the online survey, we employed the German version of the revised 20-item University of California at Los Angeles (UCLA) loneliness scale ^70,71^. Within our study, the scale displayed high internal consistency (Cronbach’s α = .91). Participants are asked to answer, how often they felt a certain way during the past two weeks, on a 4-point Likert scale with higher scores indicating more loneliness. Exemplary items are ‘I feel isolated from others.’ or ‘I do not feel alone.’ (negatively scored item). In order to assess momentary levels of loneliness in the EMA study, we used a single item measure (“Do you feel lonely at the moment?”) with a visual analogue scale (VAS; 0 -not at all, to 100 – very lonely).

#### Salivary cortisol

Saliva samples for determination of cortisol concentrations were collected at the same times as EMA. Sampling times were adapted to the individual wake-up time. Samples were taken at six time-points on two consecutive days: directly after awakening, 30 min, 45 min, 2 ½ hours and 8 hours after awakening and immediately before going to sleep. Participants stored the samples in their freezer until collected on dry ice and stored at −80°C until analysis. Analyses were conducted in the biochemical laboratory at Heidelberg University Hospital’s Institute of Medical Psychology using commercial enzyme-linked immunosorbent assay (ELISA, Demeditec Diagnostics, Germany) procedures with reported detection limit of 0.019 ng/ml. Intra-and interassay variability for cortisol were 2.95 % and 7.51% respectively. Log-transformed (ln) momentary as well as mean cortisol levels were used as outcome measures.

#### Relationship quality

Relationship quality was assessed via the short version of the *Partnerschaftsfragebogen* (PFB)^72^. It consists of 9 items that can be answered on a 4-point Likert scale. In our sample, the internal consistency of the PFB was very good (Cronbach’s α = .85). We used the global PFB score by adding up all items.

#### Control Variables

As age and sex have been previously shown to influence loneliness during the lockdown^73^, they were included as covariates into the calculations. For the EMA study, control variables (CVs) were assessed on both the momentary level (in case the outcome was cortisol) and the trait level (for both cortisol and loneliness as outcomes). Our decisions on the hormonal CVs were mainly based on expert consensus guidelines^74^. On the momentary level, the following CVs were assessed: sleep duration, sleep quality, sleeping problems, sleep medication, forced awakening, brushing teeth, eating behaviour, drinking behaviour, medication, alcohol consumption, nicotine consumption, caffeine consumption, and physical activity (with respect to the last sample), assessment time-point (1 variable for the rise from time-point 1 to 2, and 1 variable for the fall from time-point 2 to 6), and day (1 vs. 2). Trait level control variables were age, sex, and body mass index (BMI). In order to enable a trade-off between a parsimonious and a sufficiently exhausted model, we decided to include only the significant CVs at level 1. Significant CVs for cortisol as outcome were: eating, drinking, alcohol consumption, caffeine and physical activity (yes/no).

### Procedure

The study was part of a large-scale longitudinal study that aims to investigate long-term consequences of COVID-19 lockdown on psychobiological health. Results within this paper entail data from time-point 1 (first lockdown in Germany). The online survey as well as the EMA were both conducted with the platform *soscisurvey*.*de* and participation was completely anonymous. After completing the online survey, participants were asked whether they wanted to take part in the EMA. Those who were interested, were contacted via e-Mail. The responders received Salicap® tubes for saliva collection with additional informational documents via mail and specific instructions via phone. The assessment of the saliva samples took place between April 9^th^ and June 3^rd^ 2020. On two consecutive days, the participants received the respective link via SMS to a short online survey including instructions for saliva sampling six times per day. Participants were asked to refrain from food or caffeine before they provided three saliva samples which were stored in the freezer. Then, they were asked to answer further questions about their sleeping behaviour, consumption behaviour, and physical activity. Commitment was constantly monitored online: if the participants have not yet accessed the link 5 minutes after it was sent, they were reminded by phone to do so. After completion of the two sampling days, data were stored on an institute-internal data server and saliva samples remained in the participants’ home freezer until collection.

### Data processing and statistical analyses

In order to test hypotheses 1 - 3, we conducted analyses of covariance (ANCOVA). For hypothesis 1, family status (married/in a romantic relationship vs. single vs. divorced/widowed) served as independent variable (IV) and UCLA loneliness scores as dependent variable (DV). Post-hoc contrasts coding was conducted in order to analyse the linear trend of the means. For hypotheses 2 and 3, relationship status (single vs. in a relationship) and living situation (alone vs. with others) served as IVs. In this step we were interested in overall loneliness and cortisol in every-day life, thus the aggregated momentary loneliness and cortisol levels were used as DVs. As the distribution of the cortisol data was positively skewed, we natural-log-transformed the data in order to normalize their distribution. In case the assumptions of conducting an ANCOVA were violated, we used bootstrapping estimates (*n* = 1000) in order to achieve more robust results^75^. In order to test pairwise differences in momentary loneliness scores between the living situation and relationship status groups (in case the main effects were significant), we calculated Tukey Honestly Significant Differences (HSD) with *p*-values adjusted for multiple comparisons. We further calculated partial η^2^ in order to receive the effect sizes, with η^2^ ≥ 0.01 indicating a small, η^2^ ≥ 0.06 a medium, and η^2^ ≥ 0.14 a large effect.

To test hypotheses 4 and 5, we conducted multilevel modelling (MLM) regression analyses, which enabled us to assess the within- and between-person effects of momentary loneliness on momentary cortisol levels. The individual levels of loneliness were centred on the person’s mean in order to test the within-person effect on cortisol levels. In order to assess the between-person effects, we centred the individuals’ mean loneliness levels on the grand mean. For hypothesis 5, relationship status (single vs. in a relationship) and living situation (living alone vs. living with others) were included as dichotomous moderators in order to assess their interaction with level 1 loneliness scores (the exact formulas for hypotheses 4 and 5 are displayed in Appendix A in the supplement). For hypothesis 6, we conducted a multiple regression analysis with the sub-dataset of participants in a relationship, using living situation (alone vs. not alone), grand-mean-centred relationship quality (PFB) and their interaction as predictors, as well as age and sex as covariates. ANCOVA and multiple regression analyses were conducted with SPSS Statistics Version 27 ©, whereas MLM analysis were conducted via R Version 4.0.3.

## Results

In the following, we will report results from all hypotheses separately. Descriptive statistics of the outcomes of interest are shown in Table 3 and 4, respectively.

**Table 3.** Means and Standard deviations of the UCLA loneliness scale (online survey).

| Groups | Trait loneliness (UCLA loneliness scale) |  |
| --- | --- | --- |
|  | *M* | *SD* |
| Family status: |  |  |
| Married/In a relationship | 37.2 | 9.75 |
| Single | 41.09 | 11.91 |
| Divorced/Widowed | 45.42 | 12.03 |
| Sex: |  |  |
| Male | 37.18 | 10.15 |
| Female | 39.33 | 10.95 |

**Table 4.**
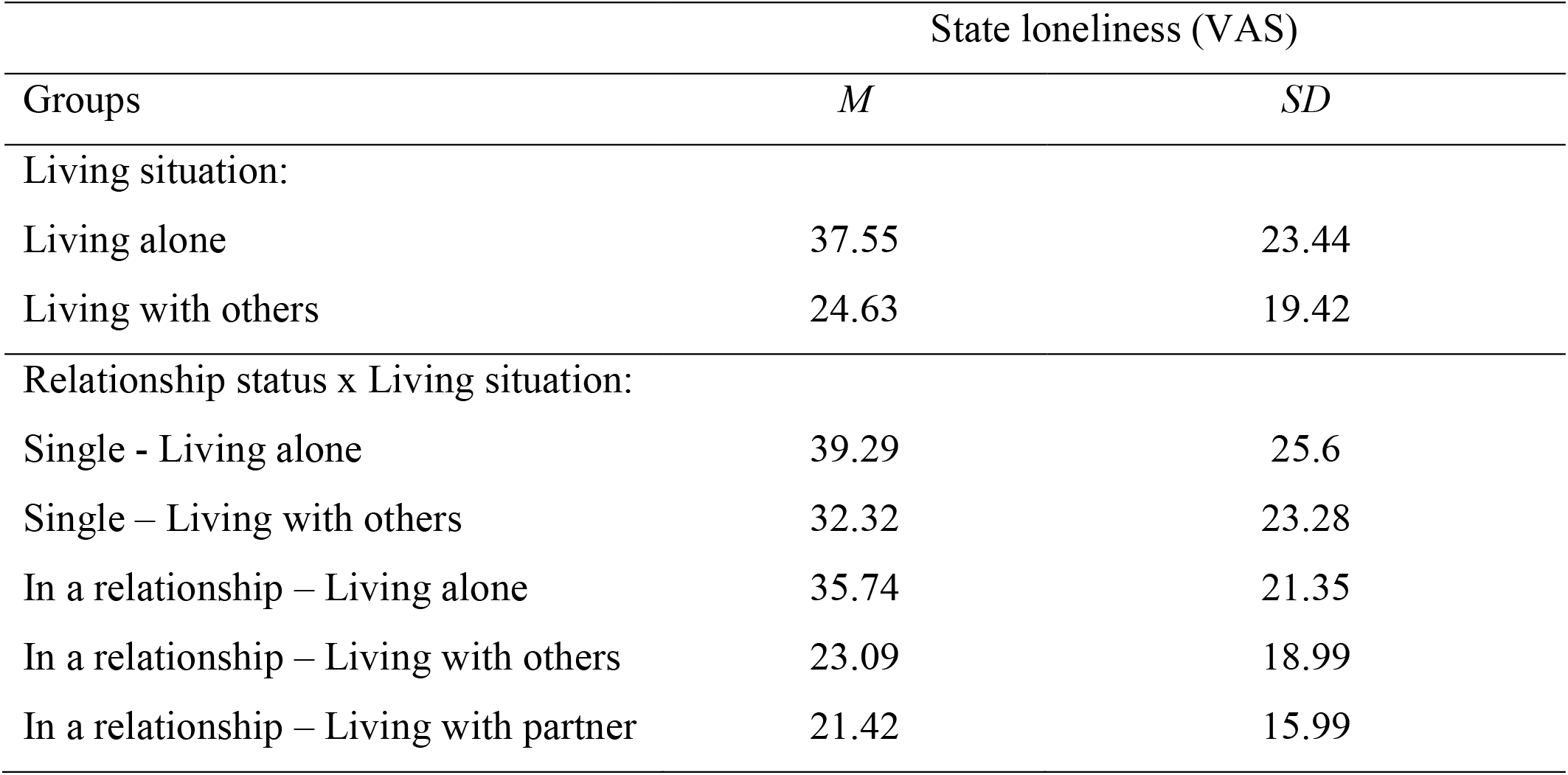
Means and Standard deviations of momentary loneliness levels (EMA study).

### Trait loneliness depending on family status (Hypothesis 1)

On average, participants had a loneliness score of *M* = 38.95 (*SD* = 10.89; *Range* = 20–77). There was a significant effect of family status on trait loneliness after controlling for sex and age (*F*(1, 1035) = 26.67, *p* < .001, η^2^ = .049). Sex was significantly related to self-reported loneliness, with women showing higher loneliness scores than men (*F*(1, 1035) = 6.39, *p* = .012, η^2^ = .006). The subsequently planned contrasts revealed a significant linear trend (*F*(2, 1035) = 26.67, *p* < .001, η^2^ = .049), indicating that married people/people in a relationship displayed the lowest loneliness scores, followed by singles and divorced/widowed individuals.

### Association of relationship status and living situation with loneliness in every-day life (Hypothesis 2)

Participants in the EMA study reported an overall loneliness of *M* = 27.36 with highly varying scores (*SD* = 20.94).

Results indicate significant associations of both living situation (*F*(1, 234) = 12.93, *p* < .001, partial η^2^ = .05) and relationship status (*F*(1, 234) = 8.57, *p* = .004, η^2^ = .04) with mean loneliness levels. People living alone reported significantly higher loneliness than people living with others. Also, individuals who were in a relationship reported significantly lower loneliness levels than singles. A third ANCOVA yielded a significant interaction between living situation and relationship status on mean loneliness (*F*(1, 233) = 7.27, *p* < .001; η^2^ = .11). Post-hoc Tukey’s HSD test indicated significant differences for the following pairwise comparisons (see Figure 2): in a relationship living alone vs. in a relationship living with partner (*p* = .016), single living with others vs. in a relationship living with partner (*p* = .028), single living alone vs. in a relationship living with partner (*p* = .001), in a relationship living alone vs. in a relationship living with others (*p* = .056), and single living alone vs. in a relationship living with others (*p* = .005).

**Figure 2.**
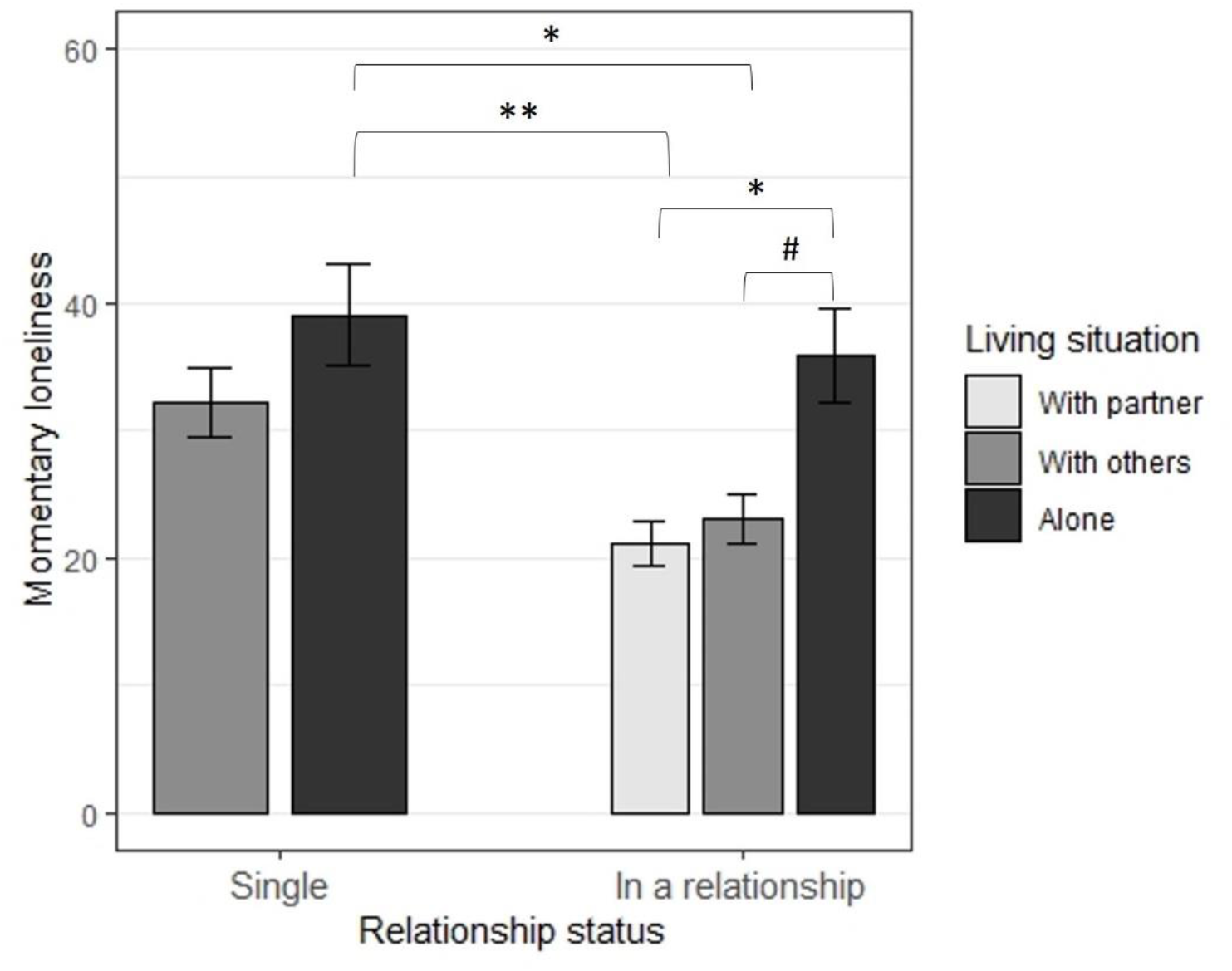
State loneliness levels (visual analogue scale) as a function of relationship status and living situation in the EMA study.

### Association of relationship status and living situation with cortisol in every-day life (Hypothesis 3)

Descriptive statistics of the variables of interest are displayed in Table 5. Mean cortisol levels in the entire EMA-sample were *M* = 8.6 ng/mL (*SD* = 2.22). Results show a significant effect of relationship status on mean cortisol levels (*F*(1, 219) = 4.58, *p* = .034, partial η^2^ *=* .02), with singles having significantly higher mean cortisol levels than individuals with a partner. Living situation did not have a significant effect on mean cortisol levels (*F*(1, 219) = 0.04, *p* = .840). Furthermore, BMI had a significant effect on cortisol, with higher BMI levels predicting higher cortisol levels (*F*(1, 219) = 15.16, *p* < .001).

**Table 5.**

| Groups | Cortisol (ng/mL) |  |
| --- | --- | --- |
|  | *M* | *SD* |
| Relationship status: |  |  |
| In a relationship | 8.44 | 6.13 |
| Single | 8.98 | 6.31 |
| Living situation: |  |  |
| Living alone | 8.64 | 2.31 |
| Living with others | 8.61 | 2.19 |

### Association of momentary loneliness, relationship status, and living situation with cortisol levels (Hypotheses 4 and 5)

The Intraclass Correlation Coefficient (*ICC*) within the empty MLM was .007, indicating that 0.7% of the variance in cortisol levels was accounted by between-person differences and 99.3% by within-person differences. As 22 cases had missing values on level 2 variables, a total of 225 cases and 1722 data points were included in the analyses. The random slopes model (with level 1-loneliness set as random predictor) showed a better fit to the data compared to the random intercepts model, (*χ*^*2*^(2) = 7.52, *p* = .020), therefore we report results from this model. There was a non-significant within-person effect of self-reported loneliness on cortisol levels (*b* =.002, *t*(1487) = 1.34, *p* = .179). Importantly, we observed a significant interaction between relationship status and momentary loneliness levels (*b* = − .004, *t*(1487) = −2.88, *p* = .004). Therefore, the association between a person’s momentary loneliness levels momentary cortisol levels was smaller for participants who were in a relationship than for those who were single. Pseudo R^2^ for this interaction was .1315, showing that the amount of unexplained variance in cortisol levels was reduced by 13.15%. The interaction between living situation and momentary loneliness levels was not significant (*b* = .002, *t*(1487) = .96, *p* = .361). Results of the entire model are shown in Tables 6 and 7 in Appendix B in the supplements.

**Table 6.** Fit indices of the multilevel model.

| Model | df | AIC | BIC | LL | L Ratio | $p$ |
| --- | --- | --- | --- | --- | --- | --- |
| 1 | 19 | 1591.11 | 1694.5 | -776.55 |  |  |
| 2 | 21 | 1587.587 | 1701.855 | -772.8 | 7.52 | .02 |

*Notes.* Model 1 = Random intercept-only model; model 2 = Random slopes-model with level 1 - loneliness set as random predictor; df = degrees of freedom; AIC = Akaike's information criterion (goodness of fit index); BIC = Schwarz's Bayesian criterion (goodness of fit index); LL = Log likelihood, L Ratio = Likelihood ratio; $p$ = $p$ -value.

**Table 7.** Results of the multilevel models with loneliness, relationship status and living situation as predictors and cortisol levels (ln-transformed) as outcome.

| Effects | Fixed Slopes Model | Random Slopes Model |
| --- | --- | --- |
| Fixed effects |  |  |
| Intercept | 2.889** (.097) | 2.896** (.096) |
| Within-person effect (L1) |  |  |
| Loneliness | 0.002<sup>#</sup> (.001) | 0.002 (.002) |
| Between-person effects (L2) |  |  |
| Loneliness | 0.0003 (.001) | 0.0004 (.001) |
| Relationship status | -0.079* (.035) | -0.078* (.035) |
| Living situation | -0.019 (.039) | -0.018 (.039) |
| Cross-Level interaction (L1xL2) |  |  |
| Relationship status*Loneliness | -0.004* (.001) | -0.004* (.002) |
| Living situation*Loneliness | 0.001 (.001) | 0.001 (.002) |
| Covariates |  |  |
| Age | 0.001 (.001) | 0.001 (.001) |
| Sex | 0.031 (.033) | 0.036 (.033) |
| Day | -0.025 (.017) | -0.026 (.017) |
| Body Mass Index (BMI) | -0.01* (.003) | -0.011* (.003) |
| Eating (yes/no) | 0.049 (.031) | 0.054<sup>#</sup> (.031) |
| Drinking (yes/no) | -0.124** (.034) | -0.130** (.034) |
| Alcohol (yes/no) | -0.048 (.038) | -0.052 (.038) |
| Caffeine (yes/no) | 0.054* (.023) | 0.053* (.023) |
| Physical activity (yes/no) | -0.004 (.022) | -0.003 (.022) |
| Time-rise<sup>a</sup> | -0.172 (.252) | -0.144 (.249) |
| Time-fall<sup>b</sup> | -0.501** (.013) | -0.501** (.013) |
| Random effects (standard deviation) |  |  |
| Intercept | .189 | .190 |
| Loneliness | - | .004 |
| Residual | .340 | .336 |

*Note.* Table depicts unstandardized coefficients (standard errors in parentheses). Number of observations = 1722; Number of participants = 225.
a -2 = time-point 1; -1 = time-point 2; 0 = time-points 3-6
b 0 = time-point 1 = time-point 4; 2 = time-point 5; 3 = time-point 6.

### Relationship satisfaction as moderator of the associations between living arrangements and loneliness (Hypothesis 6)

In the subsample of participants who were in a relationship, multiple regression analysis revealed a significant association between relationship quality and self-reported mean state loneliness levels (*t*(154) = − 2.24, ß = − .71, *p* = .026). Furthermore, participants who were living alone, showed significantly higher state loneliness levels compared to participants who were living with others (*t*(154) = − 3.13, ß = − .24, *p* = .002). However, the interaction between relationship quality and living situation was not significant (*t*(154) = 1.41, ß = .44, *p* = .162), indicating that relationship quality did not moderate the association between living situation and loneliness.

## Discussion

This study examined the (separate and joint) associations between structural (relationship status and living situation) and psychological factors (relationship quality) and loneliness and cortisol during COVID-19 lockdown.

All in all, our results provide further evidence for the belongingness-hypothesis, showing that romantic relationships, as a source for meaningful interactions and intimacy, as well as living with others protect against loneliness and neuroendocrine stress-responses, in this case diurnal cortisol levels^36-38,54,59^. Moreover, divorced/widowed participants showed the highest trait loneliness, followed by singles (never-married). Thus, the loss of previously experienced positive relationship aspects such as romantic support, solace, and physical proximity, may be associated with feelings of loneliness. Furthermore, individuals who were in a relationship and living alone (“living apart together”), were lonelier than those who were living with their partner, but did not differ in their momentary loneliness levels compared to singles living alone. Being in a relationship and living with others was associated with similar levels of loneliness compared to being single and living with others. This indicates that, during extreme physical isolation and contact restrictions, having a partner per se does not protect against loneliness, but rather living with others becomes an increasingly important buffer for loneliness. As during hard lockdown, intimacy and physical closeness are lacking in couples who are living apart, these important stress-buffering factors in the romantic relationship are suddenly missing, which is experienced as aversive^68^. Contrary to this finding, Greenfield and Russel found higher loneliness levels in couples who were living apart but with others^59^. One explanation for these conflicting findings could be that during lockdown, there were no alternatives for direct social interactions outside the apartment and thus the co-habitants became an especially important substitute for any direct contact with the romantic partner. We further found that higher relationship quality predicted lower momentary loneliness levels, which is in line with cognitive approaches to loneliness assuming that quality rather than quantity of social relationships buffers short-term psychological burden. However, relationship quality did not moderate the association between living situation and loneliness. Thus, the protective effect of living together during the COVID-19 lockdown was evident irrespectively of the relationship quality. In the online survey, female participants reported significantly higher trait loneliness levels than male participants. This adds to numerous studies revealing female gender as risk factor for loneliness^76,77^. Interestingly, however, recent neuroimaging studies indicate that loneliness-associated neural effects may be more pronounced in high lonely men than women^78,79^.

Although the results support our hypotheses about the importance of structural and psychological factors for self-reported loneliness, there are many other potential psychological mediators explaining these associations. It is important to keep in mind that romantic relationships buffer against negative mental and physical health consequences only under certain circumstances, for instance if marital functioning is perceived as positive ^33^.

Furthermore, social dimensions such as perceived social proximity, knowing that there is someone you can count on, as well as actually perceived support may be important underlying mechanisms influencing psychobiological health^29^.

On a neuroendocrine level, being in a relationship buffered momentary cortisol levels and their association with loneliness. This is in line with theoretical and empirical literature indicating that having a romantic partner serves as a biological *zeitgeber*, regulating optimal stimulation by modulating arousal levels and attenuating stress^80^. These results show us that romantic relationships have a direct impact on neuroendocrine stress responses, which in a long-term may have a positive effect on health-related outcomes^21,22^. Contrary to our hypothesis, living arrangements by themselves neither affected cortisol levels nor moderated the association between momentary loneliness and cortisol levels. One reason why we only found these associations with relationship status, could be, that there may be operators that are unique in relationships. For instance, feelings of connectedness^81^, intimacy^41^ or affective touch^82^ are specific driving factors in romantic relationships. As they are not characteristic for other relationships such as co-habitants, they only come into use when romantic relationships are investigated.

This study adds to previous research on social buffering^16,26,27,29^ in the context of enduring stress and extreme physical isolation. As lockdown-related long-term psychological health problems are increasingly revealed, it is important to study structural and psychological factors that might influence those consequences. Furthermore, short-term neuroendocrine responses during lockdown could help unravel the neurobiological mechanisms underlying detrimental effects of loneliness and social isolation for mental health. Using a psychobiological EMA design, we were able to assess not only trait loneliness levels, but also moment-to-moment variations in loneliness and salivary cortisol in a naturalistic setting. The every-day life assessments took place in the individuals’ personal environments, which yielded highly ecologically valid data. Furthermore, as the participants’ current loneliness levels were directly assessed, reporting errors due to retrospective assessment could be reduced. In order to represent the hierarchical structure of the data, MLM was used, enhancing statistical power of the analyses. Moreover, due to the close supervision of the participants, we were able to keep their commitment high and thus collect high-quality data. Another strength of this study is the wide range of the participants’ age, making the sample more representative for every age group. The collection of saliva samples in the participants’ every-day life enabled us to integrate psychobiological measures and provide a multi-level view on stress experiences during COVID-19.

This study has several limitations that need to be addressed. First of all, sample sizes differed between demographic groups. For example, 70 divorced/widowed individuals and 329 singles participated in the online survey. We recruited a convenience sample and widowers/widows and divorced individuals are on average older and less technically involved than singles, which made it more difficult to recruit them in an online survey. Another limitation is the cross-sectional design of the study, which makes it impossible to draw causal conclusions on long-term (mental) health outcomes. Furthermore, there is no baseline assessment of the variables of interest before lockdown, therefore we were not able to control for the participants’ pre-lockdown levels of loneliness and cortisol. Thus, our results can only be seen as a “snapshot” of the current situation.

There are several aspects that could be addressed in future research. Although we found main effects of relationship status, living situation, and relationship quality, they only explained a small amount of variance in the outcomes. This indicates that there are additional predictor and moderator variables influencing the outcomes. Furthermore, the stress-buffering effects of close relationships is not restricted to romantic relationships. For example, having meaningful relationships with close friends or relatives^38^ could be one protective factor. In addition, longitudinal assessments with repeated within-person measurements of loneliness and cortisol over a longer period of time could be implemented, in order to probe long-term psychological and physiological consequences of COVID-19 and strict lockdowns.

All in all, our study reveals further evidence for romantic relationships as a protective factor against trait and state loneliness, both on a structural level (alone vs. in a relationship) and a psychological level (relationship quality), as well as momentary cortisol levels during the ongoing stress of the pandemic and social isolation. Additionally, living with others during lockdown protects against loneliness in every-day life. The fact that individuals who were living apart from their partner displayed similar levels of loneliness compared to singles, implicates that especially in times of social isolation, the lack of direct physical contact to the partner makes a difference when it comes to psychological burden. This joint role of partnership and living situation should be taken into account when analysing structural factors for negative mental health outcomes, but also identifying resources for resilience. Furthermore, it is especially important to consider not only relationship status, but also relationship quality as an important psychological aspect of romantic relationships and a buffering factor for loneliness in couples, potentially counter-balancing the negative effects of living alone. This is in line with previous epidemiological research suggesting that rather than being married, it is the satisfaction with the relationship (e.g., the amount of support or criticism from a partner), which influences health-related outcomes^83^. All in all, in the context of clinical interventions, the results implicate that especially singles and divorced individuals, women, couples with low relationship quality as well as alone living residents (whether single or in a relationship) should be offered psychosocial support in order to prevent them from long-term negative health consequences. New technical methods such as smartphone apps could provide useful daily interventions or telemedical supervision. More importantly, on the one hand, individuals who are living apart from their partner, could be offered interventions to enhance their perceived relationship quality, on the other hand, alone living single individuals should be offered help in re-establishing meaningful social bonds with their close friends in order to counter-regulate their feelings of loneliness. Finally, public health campaigns should address and sensitize the society towards loneliness and mental health symptoms in those different groups to empower individuals to actively approach social offers and use them as resource.

## Data Availability

The datasets generated during and/or analysed during the current study are available from the corresponding author on reasonable request.

## Acknowledgments

We are deeply grateful for the indescribable support of C. Gäbel, who provided us her programming code for the ecological momentary assessment. We further want to thank our research assistants R. Dahlke, M. Fischer, L. Fischer, F. Frech, L.-M. Müller, N. Stockburger and J. Zimmer, without whose dedication the successful completion of the study would not have been possible. Finally, our thanks also go to the participants who made this comprehensive data evaluation possible, especially at the everyday level. This study was funded by a grant from the German Research Foundation (DFG, grant number SFB 1158) and the German Psychological Society (DGPs, Corona Scholarship). The funding sources did not have any influence on the study design, data collection, analyses or interpretation of the data, writing the manuscript, or the decision to submit this paper for publication. The authors have not been paid to write this article by a pharmaceutical company or other agency. All authors had full access to all the data in the study and had final responsibility for the decision to submit the manuscript for publication.

## Author contributions

Conceptualization: DH, ES, CAR, DS, BD, ME. Data curation: DH, ES. Formal analysis: DH, ES. Funding acquisition: DH, ES, BD, ME. Investigation: DH, ES. Methodology: DH, ES, CAR, DS, BD, ME. Resources: DH, ES, CAR, DS, BD, ME. Software: DH, ES, CAR, DS, BD, ME. Supervision: CAR, DS, BD, ME. Validation: DH, ES, CAR, DS, BD, ME. Visualization: DH, ES. Writing – original draft: DH. Writing – review and editing: ES, CAR, DS, BD, ME. The manuscript has been read and approved by all named authors.

## Figure and Tables legends

Figure 1. Participants were recruited between April 1^st^ and July 30^th^ 2020 via online media and local newspapers. Inclusion criteria were: Fluency in German, minimum age of 18 years and willingness to participate voluntarily. In total, 1483 individuals agreed to participate, from which 1054 participants filled out the questionnaires of interest.

Figure 2. Results of the Tukey’s HSD test assessing differences in mean loneliness levels of the EMA sample as a function of relationship status and living situation. ** represents *p* < .001,* represents *p* < .05, and # represents *p* < .1. Error bars depict confidence intervals based on the *t*-distribution.

## Tables

Table 1. This table depicts total and relative sample sizes split in different groups (gender, occupation and relationship status) of the Online-Study. Total *N* = 1054. Participants in the singles group are those who were never-married.

Table 2. This table depicts total and relative sample sizes split in different groups (gender, relationship status, living situation and relationship status depending on living situation) of the EMA study. Total *N* = 247. Participants in the singles group are those who were never-married.

Table 3. This table depicts means (*M*) and standard deviations (*SD*) of trait loneliness, measured by the UCLA loneliness scale, in the different subgroups of the online-study.

Table 4. This table depicts means (*M*) and standard deviations (*SD*) of momentary (state) loneliness, measured by a single-item measure with a VAS scale (0-100), in the different subgroups of the EMA study.

Table 5. This table depicts means (*M*) and standard deviations (*SD*) of momentary cortisol levels, measured by a single-item measure with a VAS scale (0-100), in the different subgroups of the EMA study.

## Appendix

### Appendix A - Formulas of the hypotheses 4 (1) and 5 (2)

1. **Level 1:** lnCort_ij_ = ß_oi_ + ß_1_ *C_lonely*_*ij*_ + ß_2_ *eat*_*ij*_ + ß_3_ *drink*_*ij*_ + ß_4_ *alcohol*_*ij*_ + ß_5_ *caffeine*_*ij*_ + ß_6_ *physical activity*_*ij*_ + ß_7_ *time_rise*_*ij*_ + ß_8_ *time_fall*_*ij*_ + ß_9_ *day*_*ij*_ + ε_ij_ **Level 2:** ß_0j_ = γ_00_ + γ_10_ *GC_lonely* + υ_0i_
2. **Level 1:** lnCort_ij_ = ß_oi_ + ß_1_ *C_lonely*_*ij*_ + ß_2_ *C_lonely*_*ij*_*\*relationship*._*i*._ + ß_3_ *C_lonely*living*_.*j*_ + ß_4_ *eat*_*ij*_ + ß_5_ *drink*_*ij*_ + ß_6_ *alcoho*l_ij_ + ß_7_ *caffeine*_*ij*_ + ß_8_ *physicalactivity*_*ij*_ + ß_9_ *time_rise*_*ij*_ + ß_10_ *time_fall*_*ij*_ + ß_11_ *day*_*ij*_ + ε_ij_ **Level 2:** ß_0j_ = γ_00_ + γ_10_ *GC_lonely*_.*j*_ + γ_11_ *age*_.*j*_ + γ_12_ *sex*._*j*_ + γ_13_ *bmi*_.*j*_ υ_0j_

Where i denotes the measurement nested in person j, vector *C_lonely* captures person-mean-centered momentary loneliness levels. *GC_lonely* captures grand-mean-centered loneliness varying on the person level (level 2), and *relationship* (0 = Single, 1 = In a relationship) and *living* (0 = Alone, 1 = With others) characteristics also varying on level 2. The vectors *C_lonely*_*ij*_*\*relationship*_*i*_ and *C_lonely*living*_.*j*_ represent cross-level interactions, with *relationship*_.*j*_ and *living*_.*j*_ being level 2 predictors. Finally, ε_ij_ denotes individual variations, whereas υ_0i_ represents differences between each person’s mean from the global mean.

Pseudo R^2^ of the significant predictors was calculated as follows: (σ^2^_0_ - σ^2^_1_)_/_ σ^2^_0_

Where σ^2^_0_ denotes the amount of variance explained before including the predictor, and σ^2^_1_ denotes the amount of variance explained after including the predictor.

### Appendix B – Results of the multilevel models

